# TPD52 promotes breast cancer cell migration, invasion and proliferation via activation of the MAPK/ERK signaling pathway

**DOI:** 10.64898/2026.08.06.26359849

**Authors:** Jiahui Yu, Zhenyu Zhu, Renhe Deng, Min Chen, Xihong Deng, JieLei Zhu, Jiankang Zhou, Xue Li

## Abstract

**Objective:** Tumor protein D52 (TPD52) is aberrantly expressed in various malignancies; however, its systematic expression profile, prognostic significance, tumor microenvironment associations, and functional mechanisms in breast cancer remain poorly defined.

**Methods:** GEO and TCGA breast cancer expression datasets were integrated to identify differentially expressed genes (DEGs). We evaluated the diagnostic performance of TPD52 via protein–protein interaction (PPI) network analysis, GO/KEGG enrichment analysis and eleven machine learning algorithms. Immunohistochemistry verified TPD52 protein expression in clinical specimens, and Kaplan–Meier analysis assessed its prognostic significance. Analysis of single-cell transcriptomic data (GSE176078) revealed the cell-type-specific distribution of TPD52 and its intercellular communication network in the breast cancer microenvironment. Weighted gene co-expression network analysis (WGCNA) explored relationships between TPD52 and tumor microbiome, hypoxia signatures as well as microsatellite instability. Moreover, TPD52 was knocked down by siRNA in MCF7 cells, and its impacts on cell migration, invasion, proliferation and the MAPK/ERK signaling pathway were examined through wound healing, Transwell, CCK-8 and Western blot assays.

**Results:** TPD52 was significantly overexpressed in breast cancer tissues at both the mRNA and protein levels. A random forest-based diagnostic model demonstrated high accuracy across multiple datasets. Kaplan–Meier analysis revealed that elevated TPD52 expression was associated with longer overall survival in specific subgroups, including the basal-like subtype, invasive lobular carcinoma, and N0/N1 stages. Single-cell analysis showed that TPD52 was predominantly expressed in tumor epithelial cells, which occupied a central position within the intercellular communication network. WGCNA further identified a positive correlation between TPD52 and a hypoxia-associated microbial module, as well as a negative correlation with a microsatellite instability module. In vitro functional assays confirmed that TPD52 knockdown significantly suppressed the migration, invasion, and proliferation of MCF7 cells, and led to reduced p-ERK1/2 protein levels.

**Conclusion:** TPD52 promotes the malignant phenotypes of breast cancer cells through activation of the MAPK/ERK signaling pathway, yet its prognostic significance is subtype-and microenvironment-dependent. These findings establish TPD52 as both a diagnostically valuable biomarker and a mechanistically defined potential therapeutic target.

## Introduction

Breast cancer remains the most prevalent malignancy among women worldwide [1, 2] and represents the leading cause of cancer-related mortality and disability-adjusted life years lost in the female population [3–5]. Global epidemiological data indicate that, in 2023, female breast cancer accounted for 2.3 million new cases (95% UI: 2.01–2.61 million) and 764,000 deaths (95% UI: 672,000–854,000), resulting in the loss of 24.1 million disability-adjusted life years (DALYs; 95% UI: 21.3–27.5 million). The Lancet projects that the global burden of breast cancer will increase substantially from 2023 to 2050, with annual new cases rising from 2.3 million to 3.56 million and deaths increasing from 764,000 to 1.37 million[6]. Despite substantial advances in surgical intervention, neoadjuvant chemotherapy, radiotherapy, and targeted therapy, the overall disease burden continues to rise persistently [7–10]. Accurate early detection and prognostic stratification are therefore essential for improving clinical outcomes.

The oncogene TPD52 is mapped to chromosome 8q21[11, 12], and members of the TPD52 family are broadly implicated in the proliferation and metastatic progression of diverse human cancers[13–18]. Emerging evidence has revealed multifaceted clinical and biological roles of TPD52 in breast cancer[19, 20]. Cheng et al. demonstrated that TPD52 is markedly upregulated in ER⁺/PR⁺/HER2⁺ breast cancer and may serve as an independent prognostic biomarker and candidate therapeutic target[21]. Using machine learning algorithms, Xia et al. systematically identified TPD52 as a promising biomarker and intervention target closely associated with immune response and clinical prognosis[20]. In triple-negative breast cancer, Shi et al. further showed that the miR-185-5p/TPD52 axis regulates tumor radiosensitivity, supporting TPD52 as a potential sensitization target for radiotherapy[22]. Notably, aberrant TPD52 overexpression is not restricted to breast cancer; it also exerts pro-tumorigenic functions in gastric cancer[17], endometrial cancer [23, 24] and lung squamous cell carcinoma [25], suggesting that TPD52 may function as a broad-spectrum tumor-associated molecule. Although prior studies have preliminarily elucidated the clinical significance of TPD52 in breast cancer and other malignancies, several critical knowledge gaps remain[13–15, 23, 26–30]. The detailed molecular mechanisms by which TPD52 drives malignant phenotypes in breast cancer are still incompletely characterized. Furthermore, the relationships between TPD52 expression and the tumor microbiota, hypoxia signatures, and microsatellite instability have not been systematically explored. Additionally, the cell-type-specific expression pattern of TPD52 within the breast cancer microenvironment currently lacks single-cell-level evidence.

In the present study, we aimed to systematically identify TPD52 as a key differentially expressed gene in breast cancer and to further explore its prognostic value, biological function, and underlying mechanisms. We first validated TPD52 overexpression across multiple public datasets and clinical specimens. Machine learning models were then constructed to evaluate its diagnostic performance, and Kaplan–Meier survival analysis was performed to assess its prognostic significance. Single-cell transcriptome analysis was employed to delineate its cell-type-specific distribution and intercellular communication patterns. WGCNA was applied to investigate the correlations of TPD52 with the tumor microbiota, hypoxia signatures, and microsatellite instability. Finally, in vitro functional assays combined with Western blotting were conducted to verify that TPD52 promotes breast cancer cell proliferation, migration, and invasion through activation of the MAPK/ERK signaling pathway. This study provides novel mechanistic insights and experimental evidence supporting TPD52 as a promising prognostic biomarker and therapeutic target in breast cancer.

## Methods

### Patients and specimens

We collected 10 tumor tissue samples from breast cancer patients who underwent surgical resection, along with 5 mammary inflammation samples for comparison, at Zhengzhou Yihe Hospital between 2023 and 2025. In this retrospective study, patients who had received antitumor therapy prior to tissue acquisition were excluded, and all participants had histologically confirmed primary breast cancer. This study was conducted in accordance with the Declaration of Helsinki. The study was approved by the Medical Ethics Committee of Zhengzhou Yihe Hospital Affiliated to Henan University (Approval No. YH-LL-KY00101). This retrospective study used archived de-identified formalin-fixed, paraffin-embedded pathological specimens obtained during routine clinical diagnosis and treatment. No additional specimens, interventions, or follow-up procedures were required. The requirement for written informed consent was waived by the ethics committee due to the retrospective nature of the study, the use of archived de-identified specimens, and the impracticability of recontacting all patients.

### Collection of BRCA data

Breast cancer expression data were obtained from the Gene Expression Omnibus (GEO) database (http://www.ncbi.nlm.nih.gov/geo), specifically from the GSE42568 series. To validate the upregulation of TPD52 mRNA expression in breast cancer tissues compared with normal breast tissues, we used the UALCAN database (http://ualcan.path.uab.edu/), an interactive web portal for in-depth analysis of TCGA gene expression data. Survival prognosis analysis and weighted gene co-expression network analysis (WGCNA) were performed using the Breast Invasive Carcinoma dataset (TCGA, PanCancer Atlas) available on cBioPortal (https://www.cbioportal.org). The single-cell sequencing dataset used for breast cancer analysis was GSE176078. The machine learning datasets comprised breast cancer and normal tissue comparisons from GSE3744, GSE42568, GSE45827, and TCGA.

### Identification of distinct gene expression patterns DEGs

The breast cancer expression dataset was downloaded from the GEO database (http://www.ncbi.nlm.nih.gov/geo) with accession number GSE42568. A total of 104 breast cancer samples and 17 normal breast biopsies were included in the analysis to identify DEGs between breast cancer and normal breast. Genes lacking corresponding gene symbols were removed, and for those with multiple probe sets, only the first name was retained. The statistical significance criteria for DEGs were set at |log2(fold change)| > 1 and p-value < 0.05. This resulted in 2,095 upregulated genes and 1,960 downregulated genes. To visualize the DEGs, a volcano plot was generated using the Weishengxin website (https://www.bioinformatics.com.cn/).

### PPI analysis and TPD52-related gene enrichment analysis

PPI network analysis was performed using the online tool STRING v12.0 (https://string-db.org/) with 2,095 upregulated genes (log2(fold change) > 1 and p-value < 0.05) in breast cancer samples as input. The PPI network was constructed with a confidence level threshold of 0.15 (low confidence), resulting in 78 TPD52-related genes. The interaction networks associated with TPD52 were visualized using Cytoscape (Version 3.10.2). To gain insights into the functional enrichment of TPD52-related genes in breast cancer, gene ontology (GO) and KEGG (Kyoto Encyclopedia of Genes and Genomes) pathway analyses were performed using the online tool DAVID v2023q1 (https://david.ncifcrf.gov/). The top 30 significantly enriched GO terms and 14 KEGG pathways were visualized using bubble plots generated with the Weishengxin website (https://www.bioinformatics.com.cn/).

### Machine learning methods

Eleven widely used machine learning algorithms were employed, including Least Absolute Shrinkage and Selection Operator (LASSO), Naive Bayes, Extreme Gradient Boosting (XGBoost), Random Forest (RF), Linear Discriminant Analysis (LDA), Ridge Regression, Generalized Linear Model Boosting (glmBoost), Support Vector Machine (SVM), Stepwise Generalized Linear Model (Stepglm), and Elastic Net (Enet). A composite approach was adopted for the final model construction. The model was trained on the TCGA dataset and validated using GSE3744, GSE42568, and GSE45827. This study developed a predictive model to systematically evaluate the disease-predictive capability of TPD52 by integrating the eleven machine learning algorithms. Model performance was assessed by calculating the area under the curve (AUC) for each model and its constituent features. Finally, the best-performing validated model was applied to each dataset individually, and model scores for each gene across datasets were obtained.

### Survival prognosis analysis

Survival data for breast cancer patients were obtained from cBioPortal (https://www.cbioportal.org/), and survival outcomes were analyzed using GraphPad Prism. Patients were stratified into low and high TPD52 expression groups using a median cut-off of 50%. Kaplan–Meier curves for overall survival (OS) were generated to illustrate the impact of TPD52 expression on patient outcomes, and the log-rank test was conducted to evaluate its prognostic significance.

### Weighted gene co-expression network analysis (WGCNA)

The relationship between the breast cancer microbiome and clinical variables was explored using WGCNA. Relevant data were obtained from the Breast Invasive Carcinoma study within the TCGA PanCancer Atlas, accessed via cBioPortal (https://www.cbioportal.org). A weighted correlation network was constructed from microbiome data using the WGCNA package in R (version 4.3.0). The optimal soft-thresholding power was determined based on the approximate scale-free topology criterion to ensure a robust network fit. After constructing the adjacency matrix with the selected threshold, microbial taxa with similar abundance patterns were grouped into modules, each containing a minimum of 15 taxa. The deepSplit parameter was set to 4, and the MEDissThres parameter was set to 0.8. The central measure of each module, termed the module eigengene, was calculated as the first principal component of the module’s abundance matrix to succinctly represent the collective expression profile of its constituent taxa. This approach facilitated the assessment of module–trait correlations, enabling the identification of modules potentially associated with TPD52 mRNA expression levels and other clinical traits. The top six microbial taxa from the most significant module correlated with TPD52 expression in breast cancer were visualized to provide insights into their relevance within the disease context.

### Single-cell RNA sequencing (scRNA-seq) analysis

Single-cell transcriptome data from breast cancer patients were acquired from the GEO database (http://www.ncbi.nlm.nih.gov/geo). This study integrated 41,515 single-cell transcriptomes from three major clinical subtypes of breast cancer, including 11 ER⁺ patients, 5 HER2⁺ patients, and 10 TNBC patients. Classical markers were used for cell annotation. scRNA-seq analysis was performed using the Seurat package (v5) in R. Low-quality cells with fewer than 200 detected genes were excluded from the analysis. The gene expression matrix was normalized using the SCTransform method, and highly variable genes (HVGs) were identified. Principal component analysis (PCA) was conducted, and the top 30 principal components were selected based on ElbowPlot and JackStraw analysis. Clustering was performed using the FindNeighbors and FindClusters functions (resolution = 2). Uniform Manifold Approximation and Projection (UMAP) was applied for visualization. Cluster-specific marker genes were identified using classical markers. When calculating the proportion of cells, the group with zero TPD52 expression was defined as low, and the group with non-zero TPD52 expression was defined as high. Cell–cell communication analysis was performed using the CellChat R package.

### Immunohistochemistry (IHC) and scoring

IHC was performed on formalin-fixed, paraffin-embedded tissue sections. Following deparaffinization, rehydration, and antigen retrieval (Tris-EDTA, pH 9.0, 100 °C, 5 min; Servicebio), endogenous peroxidase was quenched with 3% H₂O₂ (10 min). Sections were incubated overnight at 4 °C with anti-TPD52 antibody (1:200; K006300P, Solarbio), then with HRP-conjugated secondary antibody (1:1; PV-6000, Zhongshan Jinqiao) for 40 min. DAB (PV-6000) served as the chromogen, and hematoxylin as the counterstain. IHC slides were independently assessed by two blinded pathologists using the H-score (staining intensity × percentage of positive cells).

### Cell Transfection

MCF7 cells were cultured in DMEM/10% FBS at 37 °C/5% CO₂. siRNA-mediated knockdown of TPD52 (siTPD52 vs. siNC, 50 nM, Hippobio) was performed using siRNA-mate Plus (Genepharma). At 48–72 h post-transfection, migration (wound-healing inserts, KUYUAN), invasion (Transwell, LABSELECT), and proliferation (CCK-8, Abbkine) were assessed following standard protocols.

### Western blotting analysis

Protein lysates were quantified by BCA assay (Servicebio), separated by SDS-PAGE, and transferred to PVDF membranes (Cohesionbio). After blocking, membranes were probed overnight at 4 °C with primary antibodies against ERK1/2, phospho-ERK1/2 (Thr202/Tyr204), HSP90, and GAPDH (all from upingbio or Servicebio), followed by HRP-conjugated secondary antibodies for 1 h at RT. Signals were detected by ECL.

### Quantitative Real-Time PCR (qPCR)

Total RNA was extracted using RNAiso (RP101; YoungGen), and cDNA was synthesized with TransScript Uni All-in-One SuperMix (TransGen). qPCR was performed on a Bio-Rad CFX96 system using PerfectStart® Green qPCR SuperMix (TransGen) under the following conditions: 94 °C for 30 s, followed by 40 cycles of 94 °C for 5 s and 60 °C for 30 s. Relative expression was calculated via the 2⁻ΔΔCt method with GAPDH as the internal control. Primers were synthesized by Gentlegen (Suzhou, China).

### Statistical analysis

Statistical significance was assessed using Student’s t-test, the Mann–Whitney U-test, or the Chi-square test with GraphPad Prism software (version 10.0). Unless otherwise indicated, data are presented as mean ± SD, and statistical significance was defined as P < 0.05. The number of biological replicates (n) for all experiments is indicated in the figure legends. Data were visualized using R and GraphPad Prism 10.0. Clinical trial number: not applicable.

## Results

### TPD52 is identified as a key differentially expressed gene in breast cancer

This study first obtained the breast cancer expression dataset GSE42568 (104 breast cancer samples vs. 17 normal breast tissues) from the GEO database. Differentially expressed genes (DEGs) were screened using the criteria |log2(fold change)| > 1 and P < 0.05. A total of 2,095 upregulated genes and 1,960 downregulated genes were identified. The volcano plot clearly showed that TPD52 was located in the significantly upregulated region, confirming its high expression in breast cancer tissues (Fig. 1a). We further focused on 78 TPD52-centered related genes in the database, constructed a protein–protein interaction (PPI) network using the STRING database, and visualized the network with Cytoscape (Fig. 1b). Based on these 78 related genes, GO functional annotation and KEGG pathway enrichment analysis were performed using the DAVID online tool, and the results were presented as bubble plots displaying the top 30 GO terms and 14 KEGG signaling pathways (Fig. 1c). Furthermore, we analyzed the relationship between TPD52 expression levels and various clinicopathological features using the TCGA database. As shown in Fig. 1d, TPD52 expression was significantly elevated in primary tumor samples (n = 1,097) compared with normal breast tissue (n = 114; P = 1.624E−12). In the race-stratified analysis, TPD52 expression was significantly higher in tumor tissues than in normal tissues across all three subgroups: Caucasian (n = 748, P < 1E−12), African American (n = 179, P = 1.624E−12), and Asian (n = 61, P = 4.637E−09). Gender-stratified analysis revealed that TPD52 expression was significantly elevated in both male (n = 12, P = 1.74166E−03) and female patients (n = 1,075, P < 1E−12) compared with the normal group; however, given the small sample size of the male group, extrapolation of these results requires caution. Molecular subtype analysis demonstrated a significant overall difference between all subtypes and the normal group (P < 1E−12, P = 1.8511E−06, P = 4.4253E−13). Stratification by lymph node metastasis status showed that TPD52 expression was significantly higher than that in normal tissue across all N stages: N0 (n = 516, P < 1E−12), N1 (n = 362, P < 1E−12), N2 (n = 120, P = 1.624E−12), and N3 (n = 77, P = 2.723E−08). These findings suggest that TPD52 overexpression is already present prior to lymph node metastasis and persists throughout the entire metastatic process.

**Figure 1:**
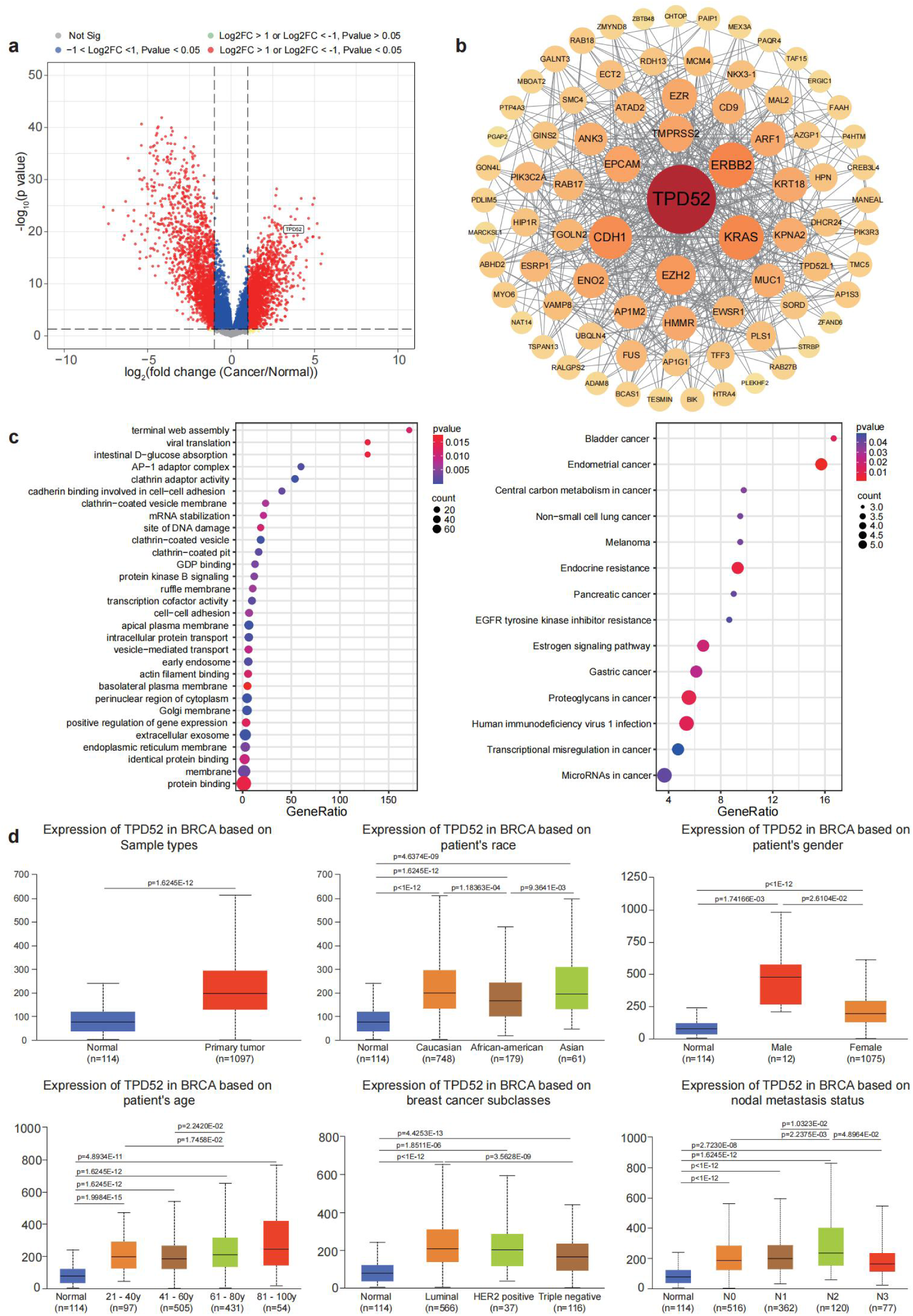
(a) Volcano plot of differentially expressed genes in the GSE42568 dataset (104 breast cancer samples vs. 17 normal breast tissues). TPD52 is located in the significantly upregulated region (|log2FC| > 1, P < 0.05). (b) Protein-protein interaction network centered on TPD52, constructed using the STRING database and visualized with Cytoscape. (c) GO and KEGG enrichment analysis of TPD52-related genes, showing the top 30 GO terms and 14 KEGG pathways. (d) TPD52 expression levels in TCGA breast cancer samples stratified by clinical features, including tumor status, race, gender, molecular subtype, and lymph node metastasis stage.

### TPD52 protein is highly expressed in breast cancer tissues

Based on the bioinformatics analyses described above, we selected TPD52 for further validation and performed immunohistochemistry on breast cancer tissues and mammary inflammation tissues (control group). The results showed that TPD52 was strongly positive in breast cancer tissues, whereas its expression level was relatively low in the control tissues (Fig. 2a, 2b). These findings confirm at the protein level that TPD52 is highly expressed in breast cancer, consistent with the transcriptomic analysis results.

**Figure 2:**
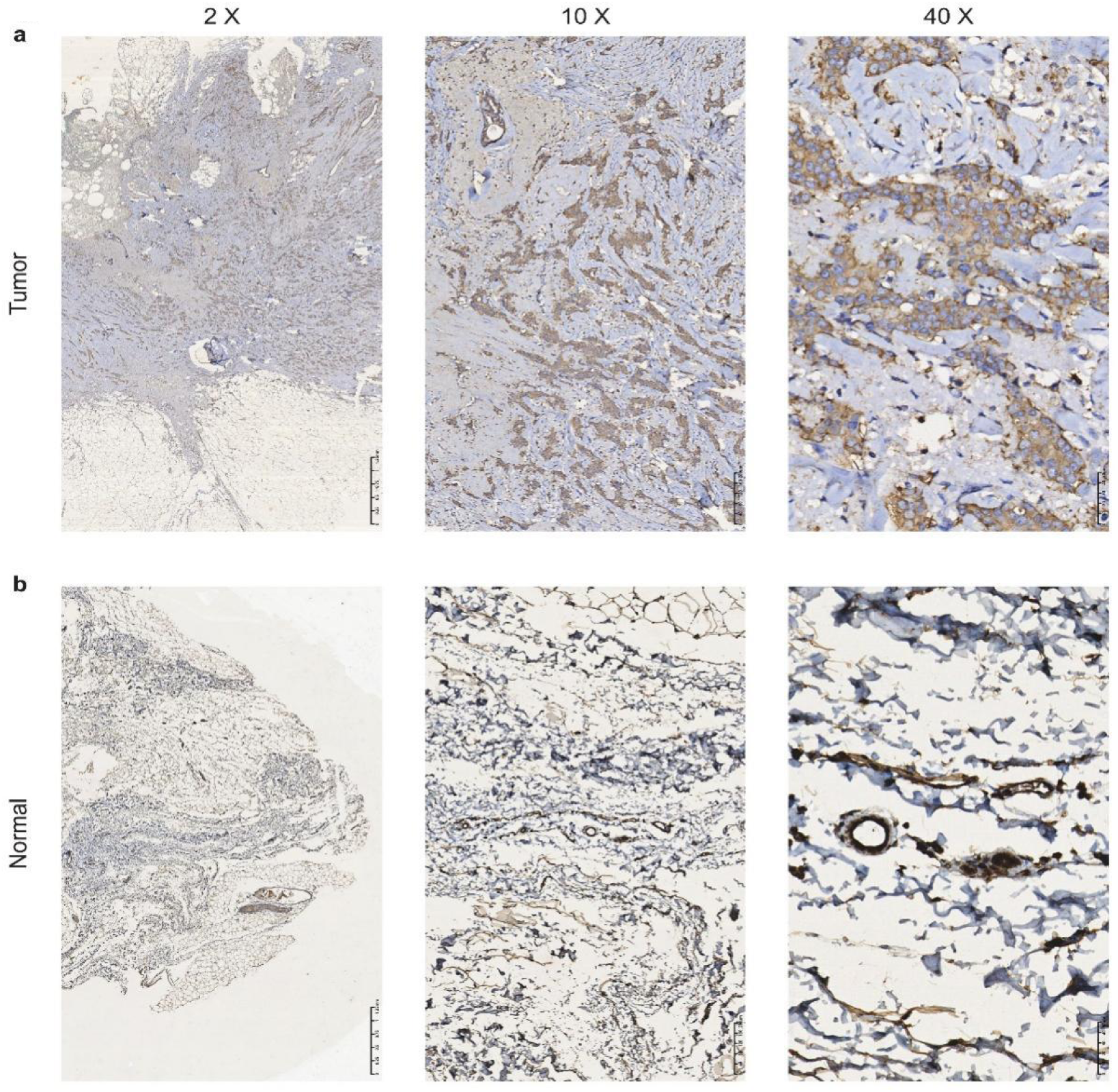
Representative immunohistochemistry images showing TPD52 expression in (a) breast cancer tissues (n = 10) and (b) mammary inflammation tissues (control, n = 5). Scale bar = 300 µm.

### Machine learning-based validation of TPD52 expression and evaluation of its diagnostic performance across multiple datasets

We systematically evaluated the diagnostic capability of TPD52 using machine learning methods in one training set and three validation sets. As shown in Figure 3a, TPD52 expression was significantly higher in breast cancer tissues than in normal tissues across all four independent datasets: GSE3744, GSE42568, GSE45827, and TCGA. Figure 3b demonstrates that the random forest model based on PPI network genes exhibited excellent predictive performance in the training set and the three independent GEO datasets (GSE3744, GSE42568, and GSE45827), with AUC values of 1.000, 1.000, 0.998, and 1.000, respectively. These results indicate that the model possesses high discriminatory ability and robust external generalizability, suggesting that the selected PPI gene signature may serve as a potential diagnostic biomarker for breast cancer. Figure 3c presents the ROC performance of TPD52 alone in each dataset, with AUC values of 0.988 (GSE3744), 0.737 (GSE42568), 0.902 (GSE45827), and 0.994 (TCGA). With the exception of a relatively low AUC in GSE42568, the AUC values in the other three datasets were all above 0.9. Overall, TPD52 can effectively distinguish breast cancer from normal tissues across different datasets, indicating that its diagnostic value is not a chance finding restricted to a single dataset.

**Figure 3:**
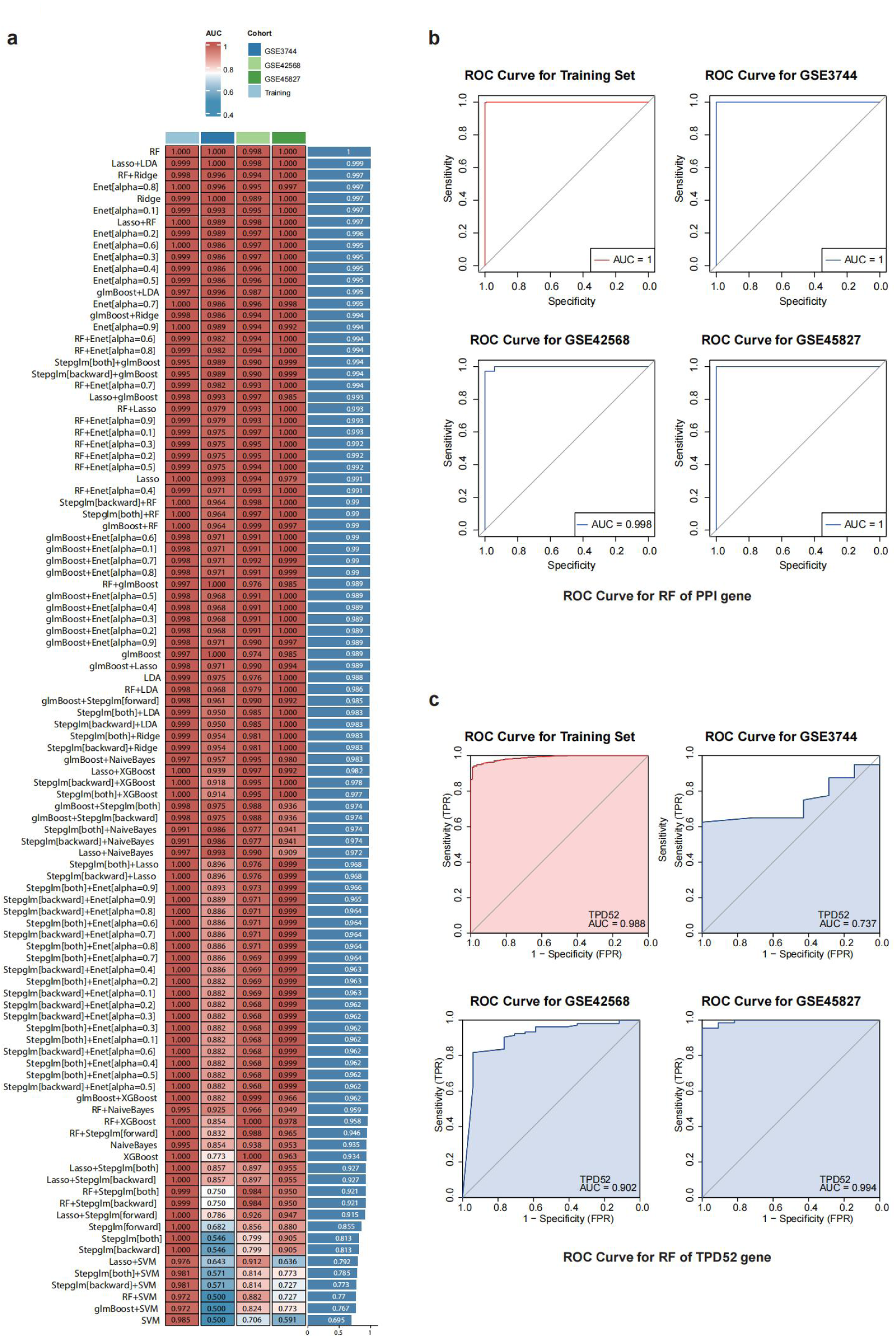
(a) TPD52 expression levels in four independent datasets (GSE3744, GSE42568, GSE45827, and TCGA). (b) ROC curves of the random forest model based on PPI network genes. (c) ROC curves of TPD52 alone as a diagnostic biomarker.

### Kaplan-Meier analysis of TPD52 expression and overall survival in breast cancer patients

To evaluate the prognostic significance of TPD52, breast cancer patients were stratified into high- and low-expression groups based on the median TPD52 mRNA level, and overall survival (OS) was compared using the log-rank test. High TPD52 expression was significantly associated with longer OS in the following subgroups: invasive lobular carcinoma (P = 0.0074, HR = 0.3034), basal-like subtype (P = 0.047, HR = 0.4168), patients aged 20–49 years (P = 0.0354, HR = 0.5026), female patients (P = 0.007, HR = 0.6412), White patients (P = 0.0477, HR = 0.6863), stage II disease (P = 0.0151, HR = 0.5429), N0/N1 lymph node status (P = 0.0047, HR = 0.5714), T3 tumor size (P = 0.0202, HR = 0.3869), and tumor-free patients (P = 0.02, HR = 0.5549) (Fig. 4), with all hazard ratios (HR) less than 1.

**Figure 4:**
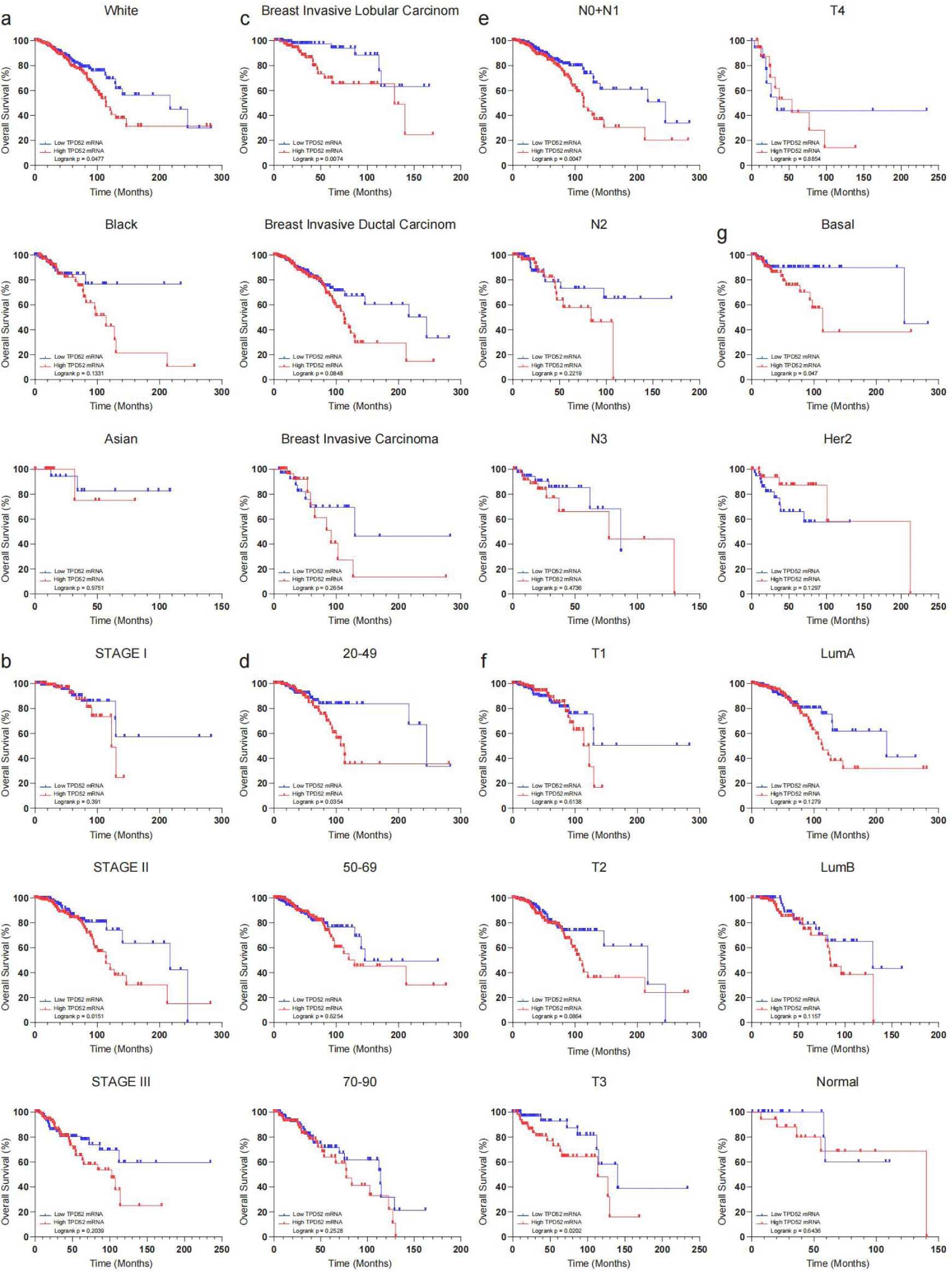
Kaplan-Meier analysis of overall survival in breast cancer patient subgroups stratified by TPD52 expression.

### Single-cell transcriptomic analysis reveals the expression pattern of TPD52 and intercellular communication in the breast cancer microenvironment

To characterize the expression pattern of TPD52 in the breast cancer microenvironment at the single-cell level, we analyzed single-cell transcriptomic data from the GSE176078 dataset, which integrates a total of 41,515 cells from 11 ER⁺, 5 HER2⁺, and 10 triple-negative breast cancer patients. UMAP clustering analysis revealed that the cells could be partitioned into multiple transcriptionally heterogeneous subpopulations (Fig. 5a). Using classical marker genes (e.g., EPCAM, CD3D, MS4A1, CD68, COL1A1, and PECAM1), these subpopulations were annotated as major cell types, including tumor epithelial cells, T cells, B cells, NK cells, plasma cells, myeloid cells, fibroblasts, and endothelial cells (Fig. 5b, 5c). The UMAP plot of TPD52 expression levels showed that high TPD52 signals were highly concentrated in the tumor epithelial cell region, in striking contrast to the immune and stromal cell regions (Fig. 5d). Cells were further stratified into TPD52 high- and low-expression groups for proportion analysis. The results demonstrated that the proportion of TPD52-high cells among tumor epithelial cells was significantly greater than that among other cell types, quantitatively confirming the predominant expression of TPD52 in breast cancer cells (Fig. 5e). CellChat-based intercellular communication network analysis revealed that tumor epithelial cells occupied a central position in the overall communication network, engaging in extensive signal exchanges with fibroblasts, myeloid cells, and other cell types (Fig. 5f, 5g). Multiple key ligand–receptor pairs were further identified (Fig. 5h–k), suggesting that TPD52-high tumor cells may participate in immune regulation and stromal remodeling of the tumor microenvironment through these signaling molecules.

**Figure 5:**
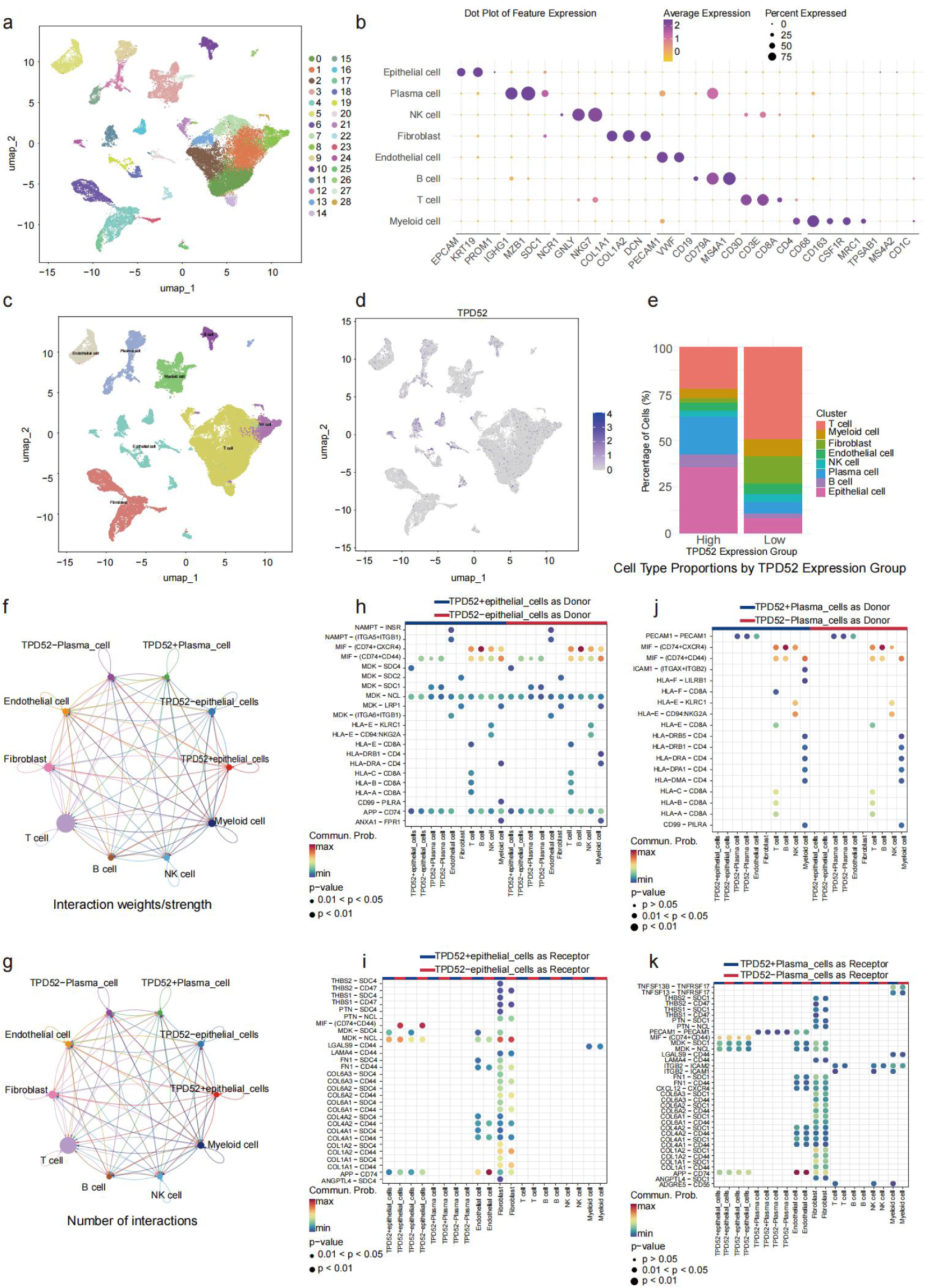
(a) UMAP clustering plot of 41,515 cells from 11 ER⁺, 5 HER2⁺, and 10 triple-negative breast cancer patients. (b) UMAP plot annotated with canonical marker genes. (c) Dot plot showing marker gene expression across different cell types. (d) UMAP plot of TPD52 expression, showing its enrichment in tumor epithelial cells. (e) Proportion of TPD52-high and TPD52-low cells across different cell types. (f, g) Cell-cell communication network plots showing tumor epithelial cells as central hubs. (h–k) Identification of key ligand-receptor pairs involved in communication between TPD52-high tumor cells and other cell types.

### Identification of breast tumor microbiota modules associated with TPD52 expression by WGCNA

To explore the potential relationship between TPD52 and the microbiota within the breast tumor microenvironment, we performed WGCNA using microbiome data from TCGA-BRCA. By constructing a weighted correlation network, microbial taxa with similar abundance patterns were grouped into distinct modules. As shown in Figure 6a, the red module exhibited a significant positive correlation with both TPD52 mRNA expression and the Ragnum hypoxia score, suggesting that the taxa within this module tend to co-vary with TPD52 and may coordinately participate in regulating the tumor hypoxic microenvironment. This implies that TPD52 may contribute to malignant progression by modulating hypoxia-related pathways. In contrast, the yellow module showed a significant negative correlation with the Ragnum hypoxia score as well as a negative correlation with TPD52 mRNA expression, indicating that the yellow module is primarily involved in hypoxia resistance and the maintenance of cellular homeostasis. As tumor hypoxia intensifies and TPD52 expression is upregulated, the overall expression of these homeostasis-protective taxa appears to be suppressed. The cyan module demonstrated significant negative correlations with both TPD52 mRNA expression and the Ragnum hypoxia score, while exhibiting a significant positive correlation with the microsatellite instability score. This suggests that the cyan module is mainly associated with DNA mismatch repair and genome stability regulatory pathways. When TPD52 expression is high and hypoxia levels are elevated, the overall abundance of this genome stability-related module is downregulated, revealing a clear inverse association between TPD52/hypoxia-related malignant phenotypes and genome stability pathways. Furthermore, we analyzed the correlation between module membership and gene significance for TPD52 expression among microbial taxa within the red module. The scatter plot revealed a significant positive correlation, indicating that microbial taxa closer to the core of this module are more strongly associated with TPD52 expression (Fig. 6b, cor = 0.74, P ≤ 0.001). Based on this, we extracted the top six microbial taxa most strongly correlated with TPD52 within the red module, including Parahospirillum, Succinimonas, Scardovia, Salinispora, Catenuloplanes, and Neorickettsia, and presented their individual correlations with TPD52 expression (Fig. 6c). This analysis identified multiple microbial species whose abundances co-vary with TPD52 expression, providing clues for future exploration of the interaction between TPD52 and the tumor microbiota.

**Figure 6:**
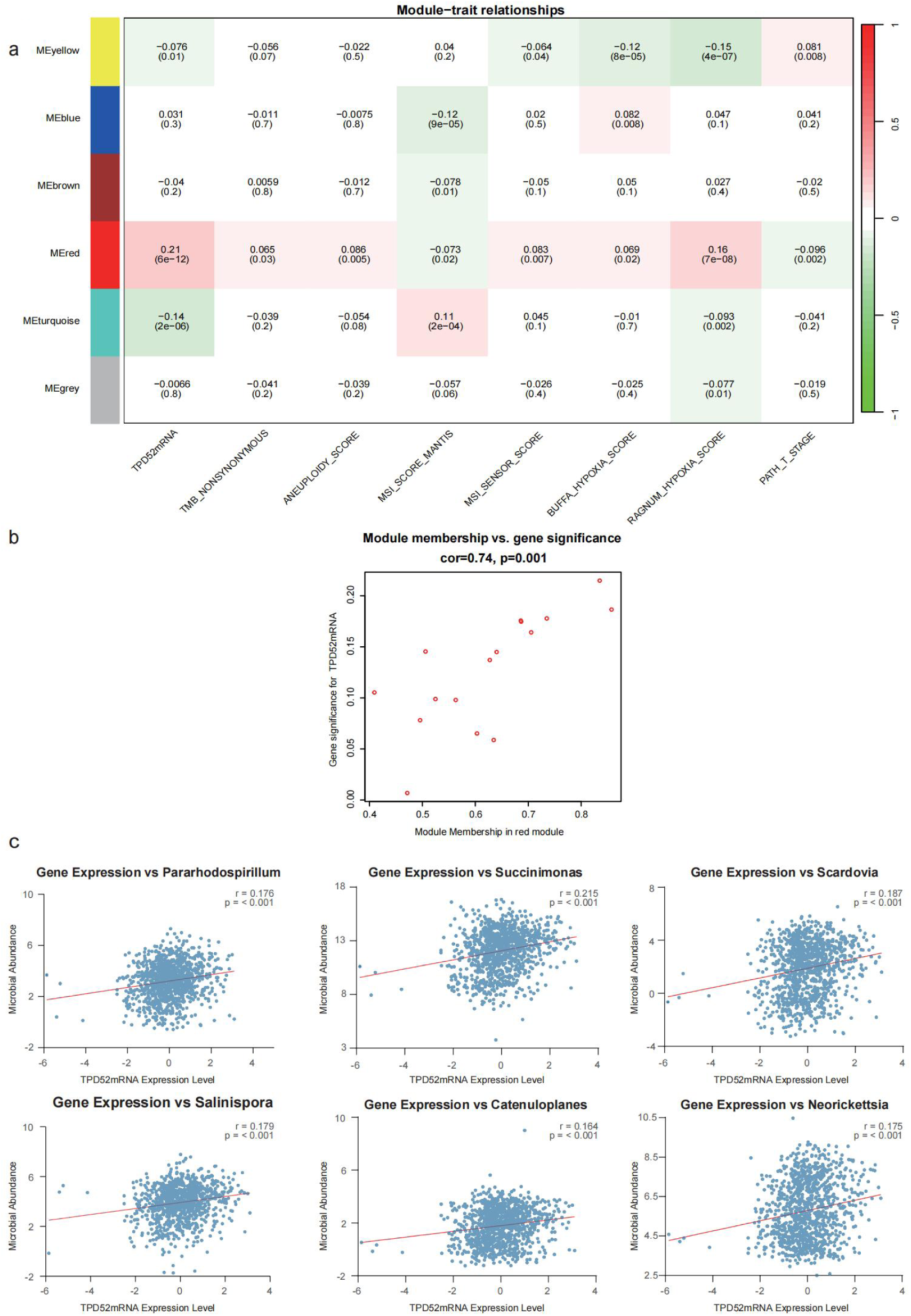
(a) Weighted gene co-expression network analysis (WGCNA). (b) Relationship between module membership and TPD52 expression. (c) Six microbial taxa associated with TPD52.

### Knockdown of TPD52 inhibits the migration, invasion, and proliferation of MCF7 breast cancer cells

To verify the function of TPD52 in breast cancer cells, we transfected MCF7 cells with siRNA to knock down TPD52 expression. Primer sequences used for RT-qPCR are provided in Supplementary Table S1, including the corresponding NCBI RefSeq accession numbers, nucleotide positions, and exon/intron locations. qPCR results showed that all three siRNAs effectively reduced TPD52 expression, with siRNA3 exhibiting the highest knockdown efficiency (Fig. 7a); therefore, siRNA3 was selected for subsequent functional experiments. Functional assays demonstrated that TPD52 knockdown significantly decreased the migratory capacity (wound healing assay, P = 0.0417; Fig. 7b, 7c), invasive capacity (Transwell assay, P = 0.0054; Fig. 7d, 7e), and proliferative capacity (CCK-8 assay; Fig. 7f) of MCF7 cells. Western blot analysis further revealed that TPD52 knockdown significantly reduced the phosphorylation level of ERK1/2 (p-ERK1/2) (P = 0.0435; Fig. 7g, 7h).

**Figure 7:**
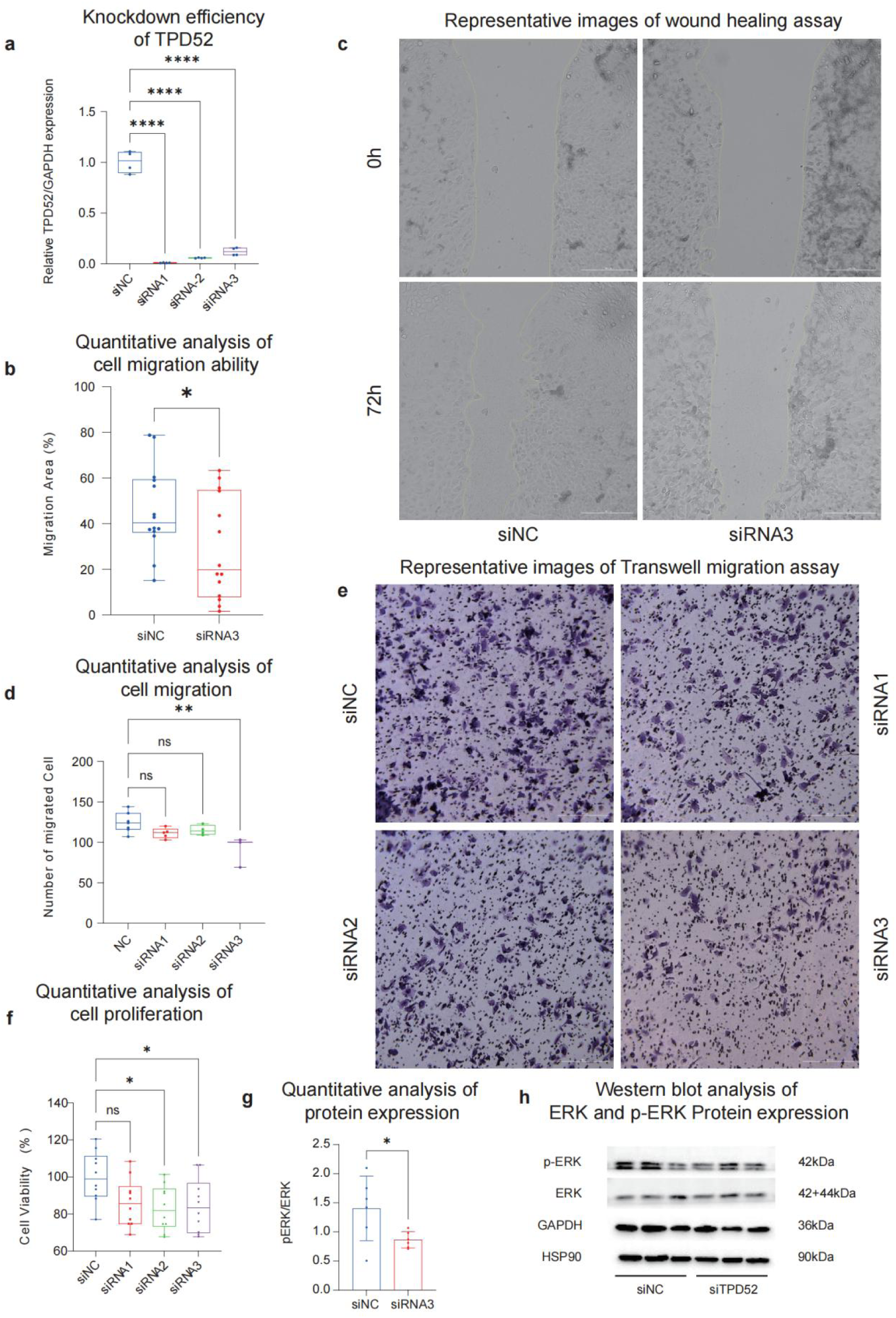
(a) qPCR assay. (b, c) Wound healing assay. (d, e) Transwell assay. (f) CCK-8 assay. (g, h) Western blot assay.

## Discussion

This study systematically investigated the expression characteristics, diagnostic and prognostic value, tumor microenvironment associations, and molecular regulatory mechanisms of TPD52 in breast cancer. By integrating public bioinformatics databases, clinical tissue specimens, machine learning models, single-cell transcriptome sequencing, weighted gene co-expression network analysis, and in vitro functional assays, we confirmed the critical role of TPD52 in the initiation and progression of breast cancer at multiple dimensions and levels. Differential gene expression analysis revealed that TPD52 was significantly upregulated in breast cancer tissues compared with normal controls, and this finding was validated at the protein level by immunohistochemistry using clinical specimens. More importantly, machine learning approaches based on four independent datasets (GSE3744, GSE42568, GSE45827, and TCGA) demonstrated that TPD52 exhibited consistently high diagnostic efficacy, achieving an AUC of 0.988 in the TCGA dataset. The random forest model constructed using TPD52-related PPI genes achieved AUC values close to or equal to 1.0 across all validation sets, underscoring its strong disease discrimination ability. These findings suggest that TPD52, either alone or as part of a multi-gene signature, holds promise as a reliable diagnostic biomarker for breast cancer. Of note, TPD52 expression was already significantly elevated in patients with N0 stage (without lymph node metastasis), indicating that its upregulation occurs at an early stage of tumorigenesis and may contribute to early detection.

To experimentally validate the functional role of TPD52 in breast cancer cells, we performed siRNA-mediated knockdown of TPD52 in MCF7 cells. Functional assays demonstrated that TPD52 knockdown significantly reduced cell migration, invasion, and proliferation, functionally confirming the driving role of TPD52 in the malignant phenotype of breast cancer cells. Western blot analysis revealed that TPD52 knockdown markedly decreased the phosphorylation level of ERK1/2, indicating that TPD52 promotes the malignant phenotype of breast cancer cells at least in part through activation of the MAPK/ERK signaling pathway. This finding is consistent with the study by Miao et al.[24] in endometrial cancer, which also demonstrated that TPD52 promotes tumor progression via the ERK/MAPK pathway, suggesting that the TPD52/ERK/MAPK axis may represent an important oncogenic signaling module across multiple cancer types. To further clarify the cellular origin of TPD52, we explored the cell-type-specific expression pattern of TPD52 within the tumor microenvironment. By analyzing single-cell RNA sequencing data from the GSE176078 dataset, we confirmed that TPD52 expression is highly concentrated in tumor epithelial cells, with only minimal expression detected in immune and stromal cells. This finding establishes that TPD52 is a driver gene originating from tumor cells themselves, rather than being passively expressed by other cells within the microenvironment. CellChat analysis further revealed that tumor epithelial cells occupy a central position in the intercellular communication network, engaging in extensive ligand–receptor interactions with fibroblasts and myeloid cells. These results suggest that TPD52-high tumor cells may actively remodel the tumor microenvironment through secreted signaling molecules. Given that TPD52 exerts pro-oncogenic functions at the cellular level, why does it appear as a favorable prognostic factor at the patient level, particularly in specific subgroups? We hypothesize that differences in the tumor microenvironment may play a key role. WGCNA based on TCGA microbiome data provided clues supporting this hypothesis. First, the red module showed a positive correlation with both TPD52 expression and the Ragnum hypoxia score, linking TPD52 to the tumor hypoxic microenvironment—hypoxia being a key driver of malignant progression in solid tumors and an important context influencing treatment response. Second, the cyan module was negatively correlated with both TPD52 expression and the hypoxia score, while positively correlated with the microsatellite instability (MSI) score. This suggests that when TPD52 expression is high and hypoxia is exacerbated, genome stability maintenance mechanisms may be suppressed. The yellow module was negatively correlated with both hypoxia and TPD52, further supporting an antagonistic relationship between TPD52-associated malignant phenotypes and protective homeostatic mechanisms. These microenvironmental regulatory insights help explain a core finding of this study: high TPD52 expression is associated with longer overall survival in specific subgroups. In recent years, multiple studies have explored the prognostic significance of TPD52 in breast cancer; however, their conclusions differ markedly from those of the present study. Cheng et al[21] identified TPD52 as an independent unfavorable prognostic factor in ER⁺/PR⁺/HER2⁺ breast cancer (HR = 1.597, P = 0.008) using multivariate Cox regression analysis of the TCGA database. Similarly, Xia and Zhou[20] identified TPD52 as an unfavorable prognostic marker in breast cancer using machine learning approaches. In triple-negative breast cancer, Shi et al[22] functionally demonstrated that TPD52 knockdown enhances radiosensitivity.

In contrast to the aforementioned studies, our findings revealed that high TPD52 expression was significantly associated with longer overall survival in subgroups such as the basal-like subtype, invasive lobular carcinoma, and N0/N1 stage, with hazard ratios below 1 in all statistically significant subgroups. We propose that this discrepancy does not reflect data inconsistency but rather highlights the context-dependent prognostic significance of TPD52. Previous studies have largely focused on ER⁺/PR⁺/HER2⁺ subtypes or mixed cohorts, whereas the favorable effect observed in our study was primarily evident in the basal-like subtype, invasive lobular carcinoma, and early-stage lymph node metastasis subgroups. These subgroups may differ in hypoxia status, MSI levels, and microbiota characteristics, thereby modulating the net effect of TPD52. In other words, TPD52 is neither a simply “good” nor “bad” gene; rather, its clinical interpretation requires stratified evaluation within specific pathological and clinical contexts.

Single-cell sequencing confirmed that TPD52 originates from tumor epithelial cells, while WGCNA further revealed that its function is modulated by microenvironmental factors such as hypoxia, microsatellite instability, and the microbiota. The MAPK/ERK signaling pathway is one of the core pathways through which cells respond to hypoxia [31], oxidative stress[32–34], and growth factor stimulation, and previous studies have demonstrated that hypoxic conditions can activate ERK1/2 phosphorylation[35–40]. In the present study, WGCNA revealed a positive correlation between TPD52 expression and the hypoxia score, while in vitro experiments confirmed that TPD52 knockdown reduces ERK1/2 phosphorylation levels. Together, these findings suggest that TPD52 may mediate the promoting effect of the hypoxic microenvironment on the malignant phenotype of breast cancer cells via the MAPK/ERK pathway, and that its net effect is not fixed but rather modulated by microenvironmental factors including hypoxia status, genomic stability, and the microbiota. The combination of these regulatory factors varies across different molecular subtypes and disease stages, which may give rise to dual—pro-oncogenic or protective—effects of TPD52. Taken together, these findings explain why TPD52 promotes malignant phenotypes at the cellular level through the MAPK/ERK pathway, yet is associated with longer survival in specific subgroups such as the basal-like subtype, invasive lobular carcinoma, and N0/N1 stage: the net effect of TPD52 depends on the molecular subtype and microenvironmental context. Furthermore, we identified the six microorganisms most strongly correlated with TPD52 expression in the red module (Parahospirillum, Succinimonas, Scardovia, Salinispora, Catenuloplanes, and Neorickettsia), offering new clues for future exploration of the “tumor gene–microbiota axis.” It is worth noting that the apparent paradox—whereby TPD52 knockdown suppresses breast cancer cell proliferation, migration, and invasion in vitro, while high TPD52 expression is associated with longer overall survival in specific patient subgroups—is not uncommon in cancer biology and can be explained by the context-dependent nature of gene function.

First, the in vitro functional assays reflect the cell-autonomous pro-malignant activity of TPD52 under optimized culture conditions, which do not recapitulate the complex tumor microenvironment (TME). In vivo, the net effect of TPD52 on patient survival may be modulated by hypoxia, immune infiltration, genomic instability, and the tumor microbiota. Our WGCNA analysis revealed that TPD52 expression is positively correlated with a hypoxia-associated microbial module (red module) and negatively correlated with modules linked to microsatellite instability (cyan module) and hypoxia resistance (yellow module). These findings suggest that TPD52-high tumors may exhibit distinct microenvironmental features—such as moderate hypoxia or specific microbiota composition—that could enhance therapeutic sensitivity or immune surveillance, thereby improving prognosis in certain contexts. Second, the favorable prognostic effect of TPD52 was observed only in specific pathological subtypes (basal-like, lobular) and early lymph node stages (N0/N1), rather than across all breast cancer patients. This subtype- and stage-restricted effect implies that the clinical outcome associated with TPD52 expression depends on the molecular background and disease progression stage. For instance, in basal-like breast cancer, which is generally more hypoxic and genomically unstable, TPD52 overexpression might be co-opted by the tumor as part of a stress-adaptive response that inadvertently limits aggressive behavior or confers vulnerability to therapy. hird, although the MAPK/ERK pathway is traditionally regarded as pro-tumorigenic, its activation can also lead to growth arrest, senescence, or enhanced drug sensitivity in specific cellular contexts, depending on the intensity and duration of signaling. Therefore, TPD52-mediated ERK activation in certain breast cancer subsets may not universally translate into a poor prognosis. Taken together, our findings do not contradict previous reports identifying TPD52 as an unfavorable prognostic factor in ER⁺/PR⁺/HER2⁺ breast cancer; rather, they extend those findings by demonstrating that the prognostic value of TPD52 is highly context-dependent. We propose that TPD52 should not be simplistically categorized as a “good” or “bad” prognostic gene; instead, its clinical interpretation requires stratification by molecular subtype, stage, and microenvironmental features.

The present study has several limitations. First, the sample size for immunohistochemical validation was relatively small (10 breast cancer tissues and 5 mammary inflammation tissues), which may have affected the specificity assessment. Second, although the favorable prognostic effect of TPD52 identified in the survival analysis is statistically robust, it still requires validation in independent cohorts with larger sample sizes and longer follow-up periods. Third, functional experiments were performed only in MCF7 cells (ER-positive, luminal subtype); whether similar effects are observed in other molecular subtypes, such as HER2-positive or triple-negative breast cancer, remains to be investigated. Finally, the mechanistic link between TPD52 and the MAPK/ERK pathway was established through knockdown experiments; however, the direct molecular interaction—for instance, whether TPD52 directly binds to components of the MAPK/ERK cascade—has not yet been elucidated.

### Conclusions

PD52 promotes breast cancer cell proliferation, migration, and invasion through activation of the MAPK/ERK pathway and exhibits high diagnostic and prognostic value, supporting its potential as a mechanistically defined therapeutic target.

## Acknowledgments

We are grateful to our peers for their suggestions and reviews of the manuscript. We would like to thank the pathologists Yang Xing and Dechen Zhang for their assistance with the IHC histological scoring. We thank all patients who contributed tissue samples and Zhengzhou Yihe Hospital Affiliated to Henan University for facilitating sample collection. We acknowledge the public databases (GEO, TCGA, cBioPortal, UALCAN, STRING, and DAVID) and the researchers who deposited the datasets used in this study.

## Declarations

### A. Author contributions

Conceptualization: Yu Jiahui; methodology: Renhe Deng; software: Yu Jiahui; validation: Zhenyu Zhu and Renhe Deng; formal analysis: Min Chen and Xihong Deng; resources: Yu Jiahui; data curation: v; writing—original draft preparation: Xue Li; Writing—review and editing: Jiankang Zhou; visualization: JieLei Zhu; supervision: Yu Jiahui; project administration: Yu Jiahui. All authors have read and agreed to the published version of the manuscript.

### B. Clinical trial number

Not applicable.

### C. Consent to participate

Not applicable. This retrospective study used archived de-identified formalin-fixed, paraffin-embedded pathological specimens obtained during routine clinical diagnosis and treatment. No additional specimens, interventions, or follow-up procedures were required. The requirement for written informed consent was waived by the Medical Ethics Committee of Zhengzhou Yihe Hospital Affiliated to Henan University (Approval No. YH-LL-KY00101) due to the retrospective nature of the study, the use of archived de-identified specimens, and the impracticability of recontacting all patients.

### D. Consent to publish

Not applicable. This manuscript does not contain any individual person’s data, images, or videos in any form that could identify a specific participant. Therefore, consent to publish is not applicable.

### F. Ethical approval

This study was conducted in accordance with the Declaration of Helsinki and was approved by the Medical Ethics Committee of Zhengzhou Yihe Hospital Affiliated to Henan University (Approval No. YH-LL-KY00101). All procedures involving human participants were performed in accordance with the relevant guidelines and regulations.

### G. Data availability statements

The datasets generated during and/or analysed during the current study are available from the corresponding author on reasonable request.

### H. ethical statement

This study was conducted in accordance with the Declaration of Helsinki and was approved by the Medical Ethics Committee of Zhengzhou Yihe Hospital Affiliated to Henan University (Approval No. YH-LL-KY00101). All procedures involving human participants were performed in accordance with the relevant guidelines and regulations.

This retrospective study used archived de-identified formalin-fixed, paraffin-embedded pathological specimens obtained during routine clinical diagnosis and treatment. No additional specimens, interventions, or follow-up procedures were required. The requirement for written informed consent was waived by the ethics committee due to the retrospective nature of the study, the use of archived de-identified specimens, and the impracticability of recontacting all patients.

